# Supervoxel-based image-to-biomarker conversions - An initial study on morphological age prediction from whole-body MRI and its clinical relevance in the UK Biobank

**DOI:** 10.1101/2025.10.24.25338723

**Authors:** Johan Öfverstedt, Alexander Blomlöf, Yasemin Utkueri, Rama K. Guggilla, Elin Lundström, Håkan Ahlström, Joel Kullberg

**Affiliations:** Department of Surgical Sciences, Uppsala University, Sweden; Department of Surgical Sciences, SciLifeLab, Uppsala University, Sweden; Antaros Medical, Mölndal, Sweden

## Abstract

Biological aging remains a central focus of research, from the scale of sub-cellular processes to whole-organism tissue morphology and function.

In this work, we developed a novel and quantitatively interpretable method for the prediction of variables, such as age, from tomographic medical images. The method uses supervoxels (whose granularity is selected by the user), standardized through inter-subject image registration, and tissue-specific feature extractions from each supervoxel to convert all image data collected into a set of well-defined imaging biomarkers. We applied the method to age prediction, using linear modelling and whole-body water-fat MRI data of 38,235 subjects in the multicentre UK Biobank study, resulting in predictions representing morphological age (MA).

We observed state-of-the-art whole-body age prediction performance on a held-out test set with a mean absolute error of 1.951/2.057 years, and R^2^ of 0.884/0.892 for females/males, respectively. The method was observed to outperform both previously reported CNN-based results from the UK Biobank and predictions from explicit biomarkers from a multi-organ/tissue segmentation approach, in a direct comparison. The interpretability of the method enabled a detailed analysis of the body-wide associations with age. Volumes of the aorta, regional muscle, bone marrow, and adipose tissue depots, and lean tissue fat content were of particular importance. The predicted MAs were of clinical relevance as they were significantly related to both type 2 diabetes and all-cause mortality. A key finding was an accuracy/utility trade-off where the more parsimonious models showed lower chronological age (CA) predictive performance but higher clinical relevance and interpretability.

The proposed method facilitates automated image-to-biomarker conversions and predictions based on subsets of anatomies, tissues, and image features, for potential application in numerous future medical studies.

This study was funded by the Swedish Heart-Lung Foundation, the Swedish Research Council, EXODIAB, and Uppsala Diabetes Center.

**Research in context:** *Evidence before this study:* We searched PubMed and Google Scholar with search terms: “uk biobank age prediction” and “whole-body mr age prediction”. Two studies involving age prediction from whole-body MR images were found, primarily using black-box AI methods, and both achieved good performance on chronological age (CA) prediction. For other imaging sequences, such as brain images, high-performance AI models have been developed. For other imaging sequences and other cohorts, age prediction using features derived from image segmentation has also been explored, commonly exhibiting lower CA prediction performance but higher interpretability.

*Added value of this study:* The study introduces a novel general-purpose image-to-biomarker method: *tissue-specific standardized supervoxel-based prediction (TS-SSP)*, relying on the definition of well-defined imaging-biomarkers through inter-subject image registration, tissue-specific supervoxel-based feature extraction, and using linear models to derive an interpretable morphological age (MA) prediction from a large dataset of whole-body MR images. The study evaluates the proposed method in comparison to segmentation-based methods, and indirectly with deep learning-based methods, and explores the interplay between model complexity, CA prediction performance, and clinical relevance. Interpretability analysis reveals spatially-resolved tissue- and organ-level associations with age and both fat content and tissue volume.

*Implications of all the available evidence:* The study shows that it is feasible to achieve age prediction with the interpretability of segmentation-based approaches with higher spatial resolution and the high CA prediction performance of deep learning-based approaches (or even higher performance) simultaneously through the use of tissue-specific standardized supervoxel-based prediction relying on image registration and linear models. The study uncovers a trade-off between the CA prediction performance and relevance to diabetes and mortality (and interpretability), underscoring the need for rigorous evaluation of age prediction methods against clinically relevant outcomes. Previously known associations with age were found, such as aorta volume and muscle volume, in addition to detailed tissue-level associations.

## Introduction

Biological age (BA) is a major risk factor for a wide spectrum of human diseases, including cancer, diabetes, and cardiovascular disorders (1). While chronological age (CA) represents the time since birth, BA reflects an individual’s physiological state, which is highly associated with chronological age but also affected by, for example, diseases, genetic factors, and lifestyle factors (26, 28). BA can be estimated through various approaches, including imaging (14), functional assessments, metabolomics, proteomics, and DNA methylation. BA has been shown to be a stronger predictor of chronic health outcomes than CA (2, 30).

Identifying MR imaging biomarkers that capture aspects of biological aging provides an opportunity for anatomically localized biomarker discovery of importance to multiple diseases. Geroprotective interventions also require proxy methods to evaluate therapy efficacy (9, 21).

The UK Biobank (UKBB) is an ongoing longitudinal study initiated in 2006 (3). Participants across the UK have contributed extensive data, including genetics, biological samples, and Magnetic Resonance (MR) images. Leveraging MR images for the non-invasive acquisition of disease-associated imaging biomarkers is vital for advancing numerous targeted prevention strategies, developing effective therapies, and improving patient outcomes (29). UKBB MR images have previously been utilized for age predictions (for varying definitions of age), where different body regions have been used as input: neck-to-knee MR images (4, 5), abdominal MR images (7, 8), and brain/cardiac MR images (6, 7, 24).

Tomographic imaging techniques collect vast amounts of information about the investigated subjects and the optimal use of this detailed information poses a significant challenge. In clinical practice, visual interpretation or simple measurements based on delineation of regions of interest are commonly used. Such delineations may be task-specific or done by more general-purpose segmentation models, such as the recent VIBESegmentator (17), which delineates 71 defined regions corresponding to various organs and tissues in whole-body MR images. Supervoxels are partitionings of 3D images based on spatial proximity and similarity of voxel intensity, providing compressed image representations. Typically, no semantic information about regions or structures of interest is required for creating the supervoxel representations, which makes them versatile and powerful. However, this lack of semantic grounding makes it challenging to relate supervoxel-based measurements between subjects. Inter-subject image registration is a process for spatial standardization of images, of which one use case is to standardize supervoxels to be mapped to well-defined anatomical regions between subjects. However, spatial standardization, which is routinely performed in some subfields, such as neuroscience, poses a challenge for whole-body images due to greater inter-subject anatomical heterogeneity compared to brain scans.

Convolutional neural networks (CNNs) have great potential in mining image data in a data-driven manner (23). Challenges, however, include an extensive need for training data, model overfitting, and shortcut learning (22, 29). Even though explainability approaches have been proposed, such as post-hoc saliency analysis, the interpretation of these models remains a significant challenge (25, 27).

The aims of this study were:

- To develop and evaluate a novel method based on tissue-specific standardized supervoxel prediction (TS-SSP) and apply it for age prediction from whole-body water-fat MR images, here referred to as morphological age (MA).
- To compare the performance of the standardized supervoxel-based features against features extracted using a state-of-the-art tissue segmentation method.
- To investigate the importance of tissue-specific feature extraction.
- To interpret the prediction models and analyze how different regions and features are associated with MA.
- Evaluate the relevance of the predicted MAs for diabetes and all-cause mortality.

## Materials and methods

An illustration of the main steps of the method is presented in Fig. 1.

**Figure 1:**
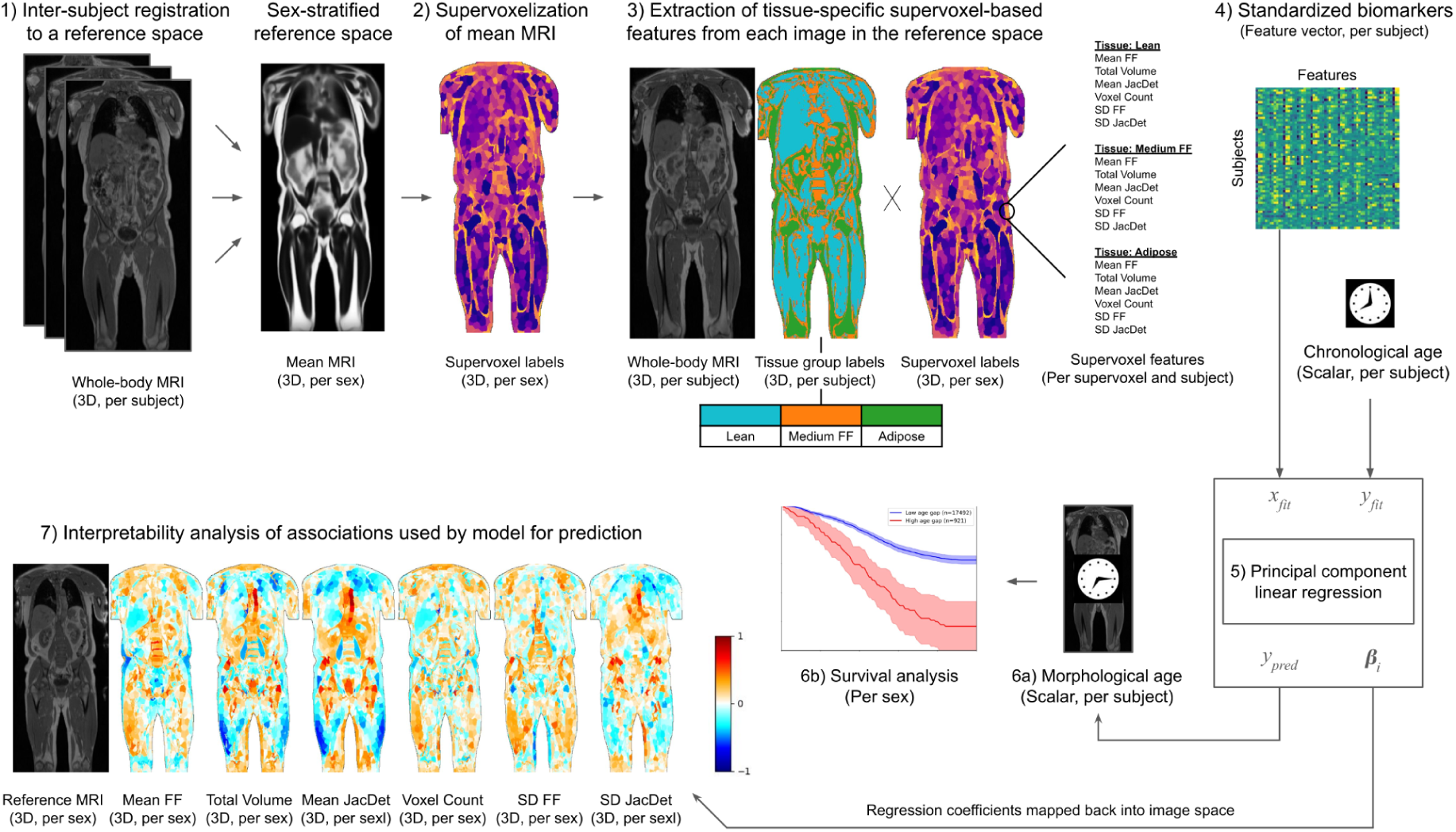
An overview of the proposed *tissue-specific standardized supervoxel prediction* (TS-SPP) method. The following steps are performed for females and males separately. 1) Whole-body Dixon MR images (with a water signal and fat signal image component from which fat fraction images are calculated) are registered and transformed into a common reference image space. 2) Supervoxels are constructed in the common image space. The supervoxels are created using a mean image calculated from all registered images. 3) Three tissue groups are defined using thresholding of fat fraction values in each original image, partitioning the voxels into groups of voxels with low, medium, and high fat content. 4) Interpretable features related to the intensities (fat fractions) and original tissue volumes (estimated from the registration through Jacobian determinants, i.e., JacDet, of the deformation fields) are calculated from within each supervoxel per tissue group (separately for the three tissue groups). The extracted data points are referred to as “supervoxel features” and can be described as extracted imaging biomarkers, all with a defined and interpretable meaning shared between all subjects and images. 5) A regression model is trained to predict a subject’s chronological age from these supervoxel features. 6a) The performance of the predicted morphological ages (MAs) is evaluated, and 6b) the association of the predicted MAs with health-related variables is evaluated. 7) Cohort-level interpretability analysis is conducted by mapping the model’s used association of each feature, tissue group (using the reference subject’s tissue group assignment for each voxel), and supervoxel combination, in the common reference image space for a sub-supervoxel visualization of the models. Image from the UK Biobank reproduced by kind permission of UK Biobank ©.

A total of 38,235 visually inspected and approved whole-body (out of 40,296, excluding 1084 due to image quality reasons, 1 due to technical reasons, and 976 due to missing CA/T2DM data), neck-to-knee MR images from the UKBB (1) were used in this study, consisting of 19,796 female subjects and 18,439 male subjects. The mean and standard deviation of age were 63.6 ± 7.4 years for females and 64.9 ± 7.6 years for males. The sex-stratified subjects were divided into ten folds per sex at random to enable cross-validation (CV), with one fold designated as the held-out test set. The test set consisted of 1,980 females and 1,844 males. The research was approved by the national ethical review authority (Dnr: 2019-03073).

The MR data were acquired using dual-echo Dixon imaging on Siemens Aera 1.5T scanners across four UK imaging centers, providing separate but inherently coregistered water signal and fat signal images. The neck-to-knee coverage was achieved using imaging across six stations, each capturing a specific region of the body. The subvolumes for both the water and fat signals from each station were stitched into a common space, resulting in 3D volumes of size 362 x 174 x 224 voxels. Fat fraction (FF) volumes were calculated by element-wise division of the fat volumes by the sum of the water and fat volumes.

Inter-subject deformable image registration to a reference image was performed to map all the images into a common space (10, 11), as seen in Fig. 1. One male and one female subject were designated reference subjects, selected based on being representative in terms of body size and body shape (both with CA≈64 years). The registration (15) was performed on water and fat fraction volumes, along with subcutaneous fat and muscle masks generated by VIBESegmentator (17), applied on the sum of the water and fat images. For each deformation field, a deformed FF image was computed along with the Jacobian determinant (JacDet) field representing the local estimated reference voxel-wise expansion or contraction. More details are presented in Supplementary A.

A sex-stratified mean FF volume was created and used as input to generate a supervoxelization (to reduce bias, compared to using a single image) with the Simple Linear Iterative Clustering (SLIC) algorithm (13), which generates a configurable number of supervoxels *n_seg_* (more details are found in Supplementary B). Figure 2 shows the mean FF volumes and supervoxelizations for various choices of the number of supervoxels.

**Figure 2.**
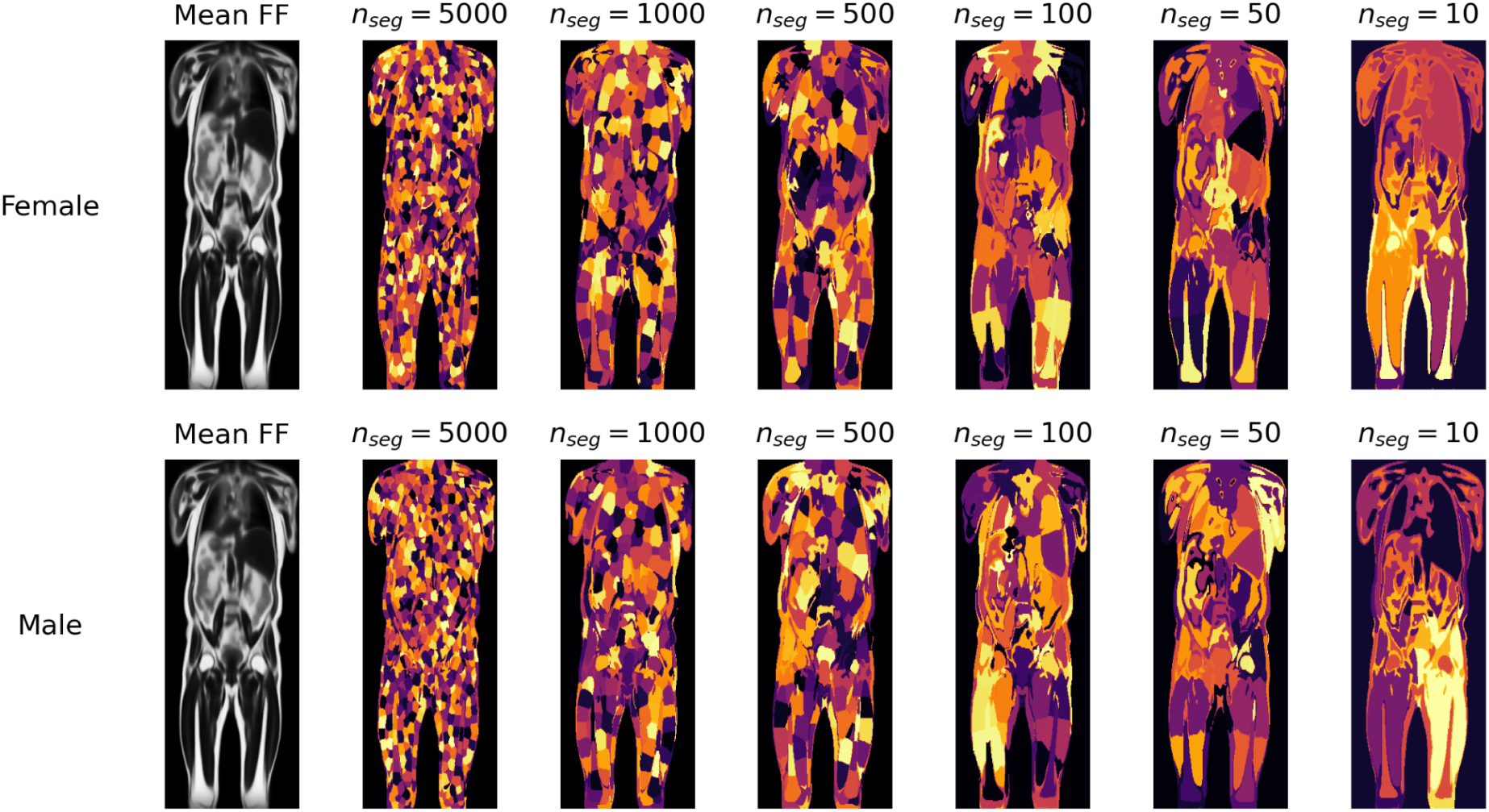
Visualization of the mean image for each sex on the left, and different supervoxelizations on the right corresponding to varying numbers of supervoxels. A higher number of supervoxels (parameterized by *n_seg_*) provides a more detailed partitioning of the body, a larger number of features, each of which is computed from a smaller number of voxels. Image from the UK Biobank reproduced by kind permission of UK Biobank ©.

For each subject, we extracted features from the voxels within each standardized supervoxel (20). To alleviate partial volume effects resulting from the presence of multiple tissue types (e.g., both adipose tissue and lean tissue) within a single supervoxel, we propose extracting features specific to user-defined tissue groups. The tissue-specific features were selected based on CV performance. These were: mean FF, total volume (i.e., mean JacDet*voxel count), mean JacDet, reference space voxel count, standard deviation (SD) FF, and SD JacDet. See detailed definitions in Supplementary C.

We defined three tissue groups based on FF, here denoted “low FF” (lean tissue), “medium FF”, and “high FF” (adipose tissue), for a method referred to as the *3-tissue group configuration*. These thresholds were selected empirically based on the histogram of FF values. Denoting *x* a voxel, *I(x)* the FF of voxel *x*, and *T***_***(x)* tissue group weights, these 3 tissue groups were defined as binary categorizations: *T***_low_***(x) = 1* if *I(x) ≤ 20%, 0 otherwise, T***_high_***(x) = 1* if *I(x) > 80%, 0 otherwise,* and *T***_medium_***(x) =* 1 **-** *T***_low_***(x) - T***_high_***(x)*. For comparison, we also consider a version of the method where all the voxels were assigned to a single tissue group (*T***_all_***(x)*=*1*), referred to as the *1-tissue group configuration*, equivalent to feature extraction without tissue groups.

We applied Z-score normalization with clipping (below −3 SD, above 3 SD) to the features (20). Principal component analysis (PCA) dimensionality reduction with 3000 components was applied to the z-scores to combine correlated features into components (20), and to reduce the risk of shortcut learning (22). Additionally, we selected a subset of 10 out of 3000 PCA components, maximizing the univariate absolute correlation with CA. These operations were performed fold-wise based on training set statistics. Linear models, selected for both simplicity and interpretability, were fitted to predict the CA from the PCA components using 9-fold CV. To visualize the used associations and interpret the predictions, the regression coefficients were mapped to each supervoxel/tissue/feature combination z-score, normalized through division by the maximum absolute value, and plotted in the reference space.

The code for the method is shared as open source (19).

### Evaluation and statistics

The performance of the prediction models was evaluated on both the validation data and on the held-out test set, with the CA as reference. The measures used to quantify the prediction errors were 1) the mean absolute error (MAE), which decreases with improved performance, and 2) the coefficient of determination *R^2^*, which increases with improved performance. Ablation studies were conducted to examine how age predictions were influenced by training set size, number of supervoxels, the impact of the 3-tissue group configuration compared to the 1-tissue group configuration, as well as tissue-specific experiments where age was predicted from features extracted from a 1-tissue group and from a single tissue group and feature combination.

To evaluate the advantage of the standardized supervoxel features in comparison to alternative available segmentation approaches, we also considered a recently published method, VIBESegmentator, which segments whole-body MRI scans into 71 regions comprising organs and tissues. From these segmentations and tissue-group masks, we extracted features analogously to the supervoxel-based features.

We performed statistical tests to compare the absolute errors of the 3-tissue group configuration (TS-SSP) compared to the 1-tissue group configuration, both using supervoxels, as well as the 3-tissue group supervoxel method compared to the 3-tissue group VIBESegmentator method. 36 tests (9 per sex and method) were performed; therefore, Bonferroni correction with a factor of 36 was applied to adjust the p-values.

To evaluate the individual importance of each tissue and feature, tissue- and feature-wise predictions were performed.

Interpretability analysis was performed on both the 10-component and 3000-component models by reading the 3D interpretability maps (30) by a radiologist with 40 years of experience.

In addition to the evaluation of the age predictions, we investigated the relevance of the predicted ages to the prevalence of type-2 diabetes (T2DM) and all-cause mortality. After bias correction (described in detail in Supplementary G), which removes systematic underestimation of high ages and overestimation of low ages (24), we computed an age gap as the difference between each bias-corrected predicted age and the subject’s CA (age gap = MA-CA).

We plotted a moving average of the age gaps for the subjects with T2DM as a function of CA. Bootstrapping (with 1000 repetitions) was applied to the moving averages to obtain 95% confidence intervals (CI) of the average age gaps. We also performed logistic regression with either CA or MA as input, and compared the AUC values using DeLong’s test with a Bonferroni correction factor of 2.

The survival analysis consisted of Kaplan-Meier plots with two groups: 1) subjects with an age gap larger than the top 10th percentile, and 2) remaining subjects. Groupwise bootstrapping (with 10000 repetitions) was applied to the two groups to obtain a 95% CI. Statistical testing with the log-rank test was performed with a Bonferroni correction factor of 2. For comparison of the CA and MA as predictors of mortality, we also performed logistic regression to predict 5-year all-cause mortality with either CA or MA as independent variables, and the resulting area under the curve (AUC) values were compared with DeLong’s test with a Bonferroni correction factor of 2.

## Results

The performance of the proposed method in comparison to the reference CAs is presented in Table 1, where the 3-tissue group configuration significantly outperformed the 1-tissue group configuration in terms of MAE and R^2^, both on the cross-validation experiments and on the test set, except for fold 6 in males (p-values are shown in Supplementary H).

**Table 1:** The performance of the two primary models (with 3000 PCA components and 5000 supervoxels) in cross-validation (CV) experiments and on the test set, which consists of 1,980 samples in the female cohort and 1,844 samples in the male cohort. Performance was measured using R^2^ and MAE, with CA as reference. Each cell presents the mean results of nine models on the respective partition, fitted using cross-validation, with the standard deviation across models in parentheses.

| Tissue groups | Partition | $R^2$<br>Female | $R^2$<br>Male | MAE (years)<br>Female | MAE (years)<br>Male |
| --- | --- | --- | --- | --- | --- |
| 1-tissue group | CV | 0.868<br>(0.004) | 0.871<br>(0.005) | 2.131<br>(0.026) | 2.179<br>(0.024) |
| 3-tissue group<br>[TS-SSP] | CV | **0.882**<br>**(0.004)** | **0.886**<br>**(0.003)** | **2.006**<br>**(0.028)** | **2.043**<br>**(0.022)** |
| 1-tissue group | Test set | 0.869<br>(0.001) | 0.879<br>(0.001) | 2.074<br>(0.006) | 2.166<br>(0.011) |
| 3-tissue group<br>[TS-SSP] | Test set | **0.884**<br>**(0.001)** | **0.892**<br>**(0.001)** | **1.951**<br>**(0.009)** | **2.057**<br>**(0.014)** |

The cohort-level interpretability analysis for the 10-component (exhibiting strong association to disease and survival) and 3000-component (exhibiting strong performance on CA prediction) 3-tissue group configuration of the method is visualized in Fig. 3 and Fig. 4. See also movies of the full 3D results in (16).

**Figure 3:**
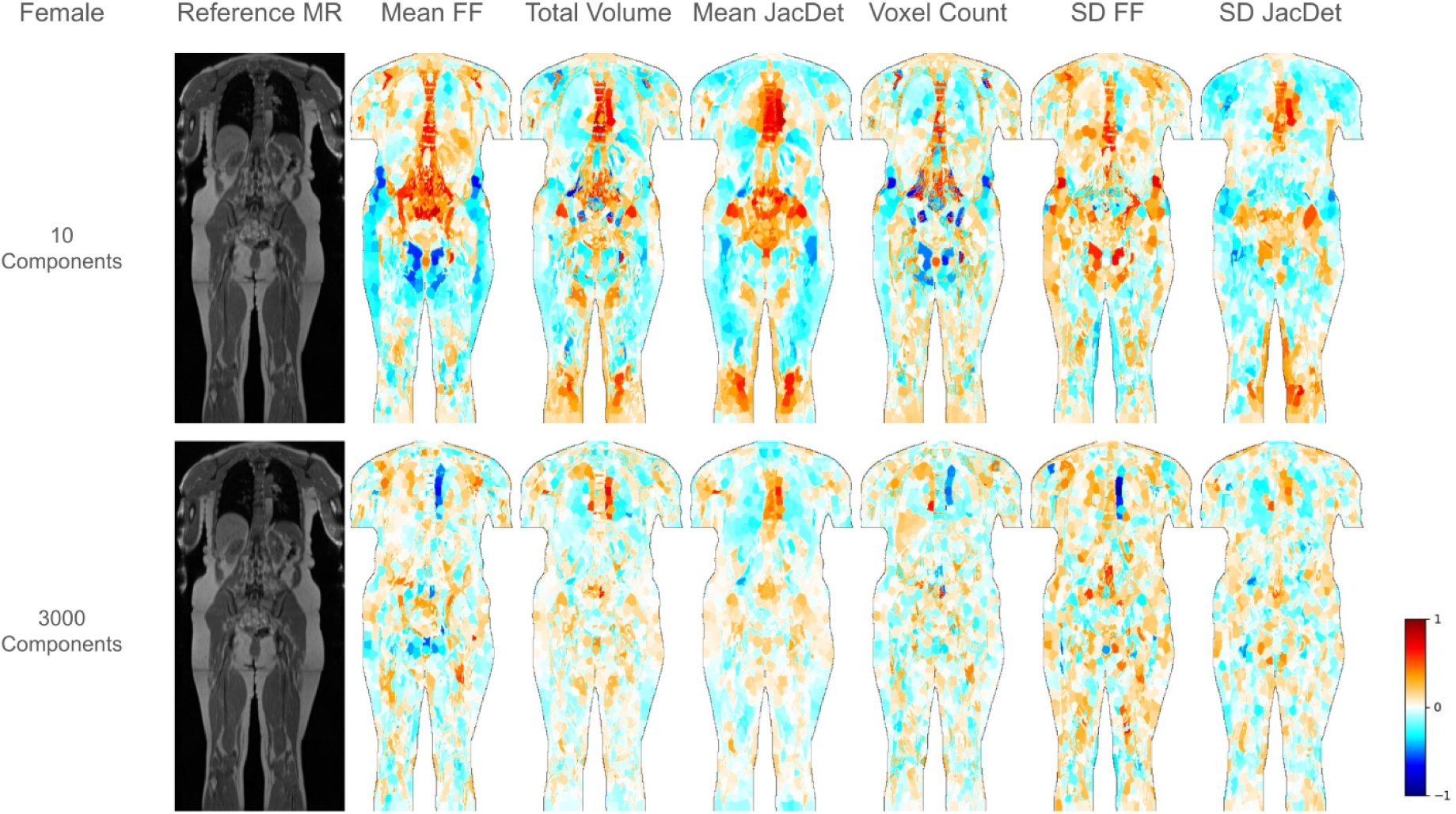
Cohort-level interpretability analysis for the female subjects. The leftmost column of each subfigure shows a slice from the reference volume, and the six subsequent images on each row show the association used by the models. The associations are normalized and expressed in standard deviations of each supervoxel feature. The tissue group information within each supervoxel in the reference subject is used to visualize the results on a sub-supervoxel level. The 10-component models exhibit a strong association with disease and survival (shown in Fig. 6 and Fig. 7), and the 3000-component models exhibit strong performance on CA prediction (shown in Tab. 1). Image from the UK Biobank reproduced by kind permission of UK Biobank ©.

**Figure 4:**
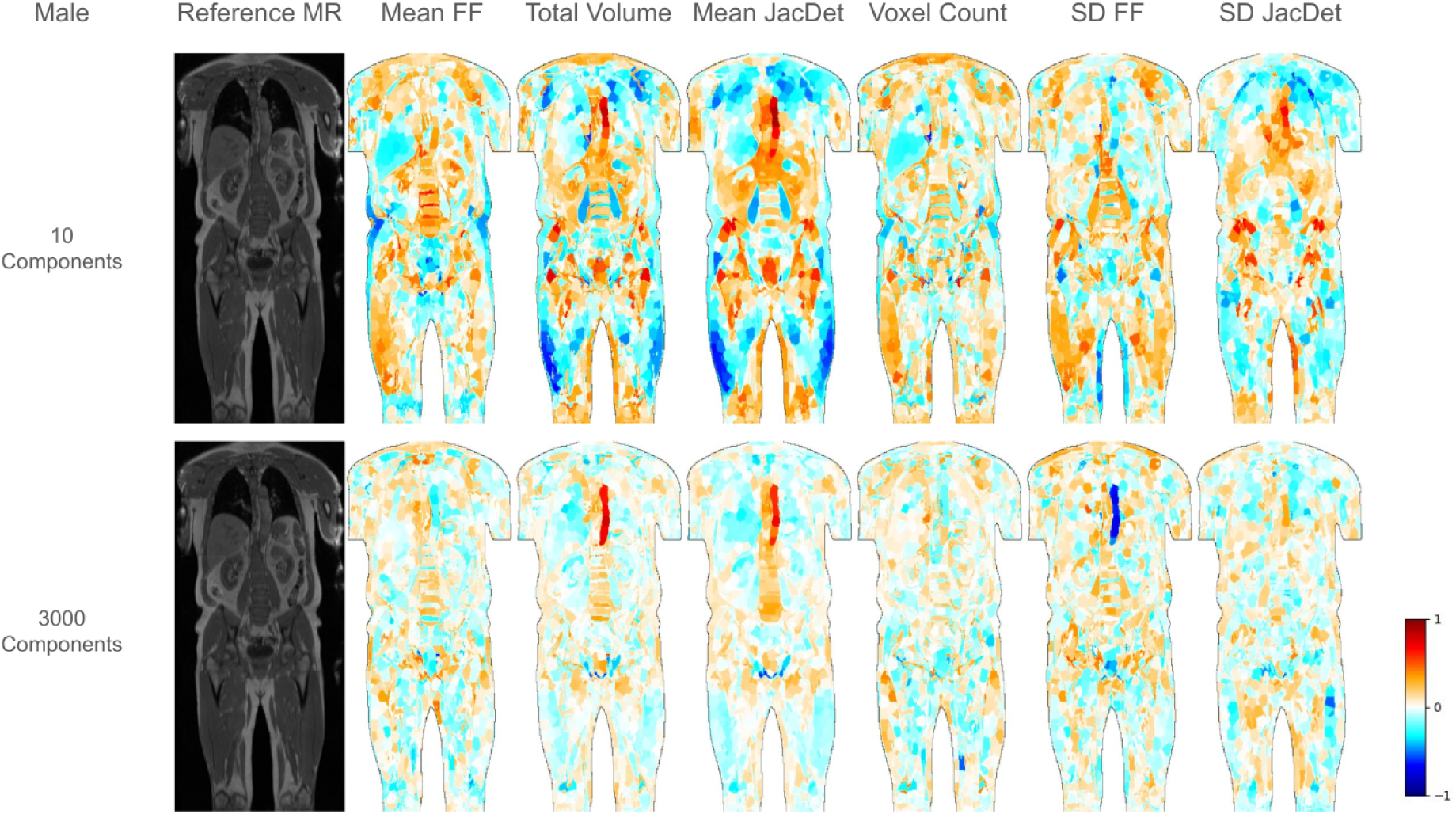
Cohort-level interpretability analysis for the male subjects. The leftmost column of each subfigure shows a slice from the reference volume, and the six subsequent images on each row show the association used by the models. The associations are normalized and expressed in standard deviations of each supervoxel feature. The tissue group information within each supervoxel in the reference subject is used to visualize the results on a sub-supervoxel level. The 10-component models exhibit a strong association with disease and survival (shown in Fig. 6 and Fig. 7), and the 3000-component models exhibit strong performance on CA prediction (shown in Tab. 1). Image from the UK Biobank reproduced by kind permission of UK Biobank ©.

For the 10-component models, we observed a positive model association between MA and the volume of the aorta, prostate (in males), visceral adipose tissue (VAT), vertebrae, bone marrow, pulmonary artery (at the bifurcation), gluteus maximus (more distinctly in females), and lower ventral abdominal SAT (more distinctly in females). We observed a negative model association between age and the volume of the liver, spleen, lungs, pancreas (more distinctly in females), kidneys, heart ventricles, upper ventral abdominal SAT, muscles of the neck, back, and the thighs, pulmonary artery (at the origin), femurs (predominately negative) (Fig. 3 and 4). We observed a positive model association between age and mean FF of the kidneys, spleen, lower ventral abdominal SAT, pancreas, vertebrae, bone marrow, humerus, femur (weak), and most muscles (distinctly in the back muscles). We observed a negative model association between age and the mean FF of the liver, VAT, dorsal and side abdominal SAT, and (predominantly negative) leg SAT (Figs. 3 and 4). Comparing the 10- and 3000-component models, the 10-component models exhibit anatomically coarse associations, and the 3000-component models exhibit more anatomically detailed associations, with more intra-tissue and intra-organ variations. Several regions exhibit a change of sign of the association between the configurations. The volume of the aorta was a key feature in both configurations for both sexes (Figs 3 and 4).

The impact of the number of supervoxels and dataset size on age prediction performance is shown in Figure 5, and in Supplementary I, respectively, where more supervoxels and more data led to improved performance.

**Figure 5:**
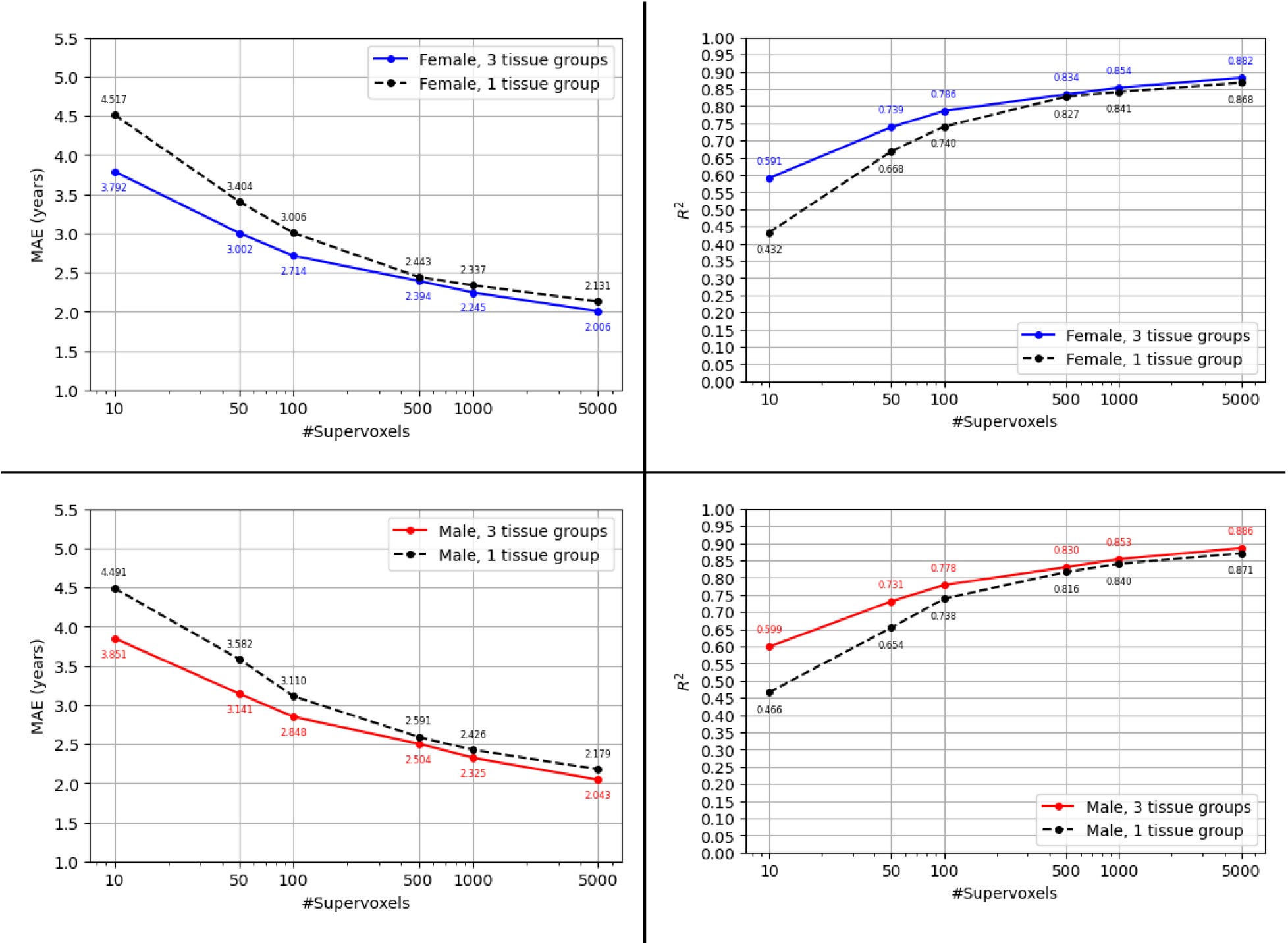
The effect of different numbers of supervoxels on MAE and R^2^ in the cross-validation (CV) evaluation of chronological age (CA) prediction (evaluated on the CV sets), with a comparison of the 1-tissue group configuration with the 3-tissue group configuration. A lower MAE and a higher R^2^ indicate better performance. The y-axis is scaled linearly, while the x-axis is scaled logarithmically.

The performance comparison between supervoxel features and VIBESegmentator-based features is shown in Table 2, where the supervoxel-based predictions outperform the segmentation-based predictions. We observed that the 3-tissue-group feature extraction also aided the performance of the VIBESegmentator-based age predictions. The p-values are shown in Supplementary H.

**Table 2:** Performance comparison between the main proposed models, which perform predictions using features extracted from 5000 supervoxels, and a model that performs predictions using features extracted from masks produced by VIBESegmentator. The performance was measured using R^2^ and MAE. Each cell presents the mean results of nine cross-validation models on the test set, with the standard deviation across models in parentheses.

| *Tissue groups* | *Regions* | *$R^2$*<br>*Female* | *$R^2$*<br>*Male* | *MAE (years)*<br>*Female* | *MAE (years)*<br>*Male* |
| --- | --- | --- | --- | --- | --- |
| 1-tissue group | Standardized supervoxels | 0.869<br>(0.001) | 0.879<br>(0.001) | 2.074<br>(0.006) | 2.166<br>(0.011) |
| 1-tissue group | VIBESegmentator | 0.676<br>(0.001) | 0.720<br>(0.001) | 3.302<br>(0.004) | 3.257<br>(0.003) |
| 3-tissue groups<br>[TS-SSP] | Standardized<br>supervoxels | **0.884**<br>**(0.001)** | **0.892**<br>**(0.001)** | **1.951**<br>**(0.009)** | **2.057**<br>**(0.014)** |
| 3-tissue groups | VIBESegmentator | 0.712<br>(0.001) | 0.764<br>(0.001) | 3.115<br>(0.008) | 3.035<br>(0.009) |

35 tests out of the 36 were statistically significant in the comparisons between 3-tissue groups and 1-tissue group, and supervoxel-based features and VIBESegmentator-based features. The 3-tissue group configuration was superior to the 1-tissue group configuration except for male fold 6, and 3-tissue-group supervoxel-based features outperformed 3-tissue-group VIBESegmentator features.

Results from the tissue- and feature-wise age predictions are listed in the supplementary M. The numerically best performing predictions were found to be voxel count in medium FF tissue, and the weakest was SD JacDet in adipose tissue.

The results of the association of the age gap and prevalence of T2DM are presented in Fig. 6, where we observed a substantial positive age gap in the subjects with T2DM compared to the control group. Prediction of T2DM was found to improve by using MA instead of CA. Logistic regression yielded an AUC delta of 5.5% for males (63.4% compared to 57.9%), p=0.0000, and 1.4% for females (63.0% compared to 61.6%), p=0.0475.

**Figure 6:**
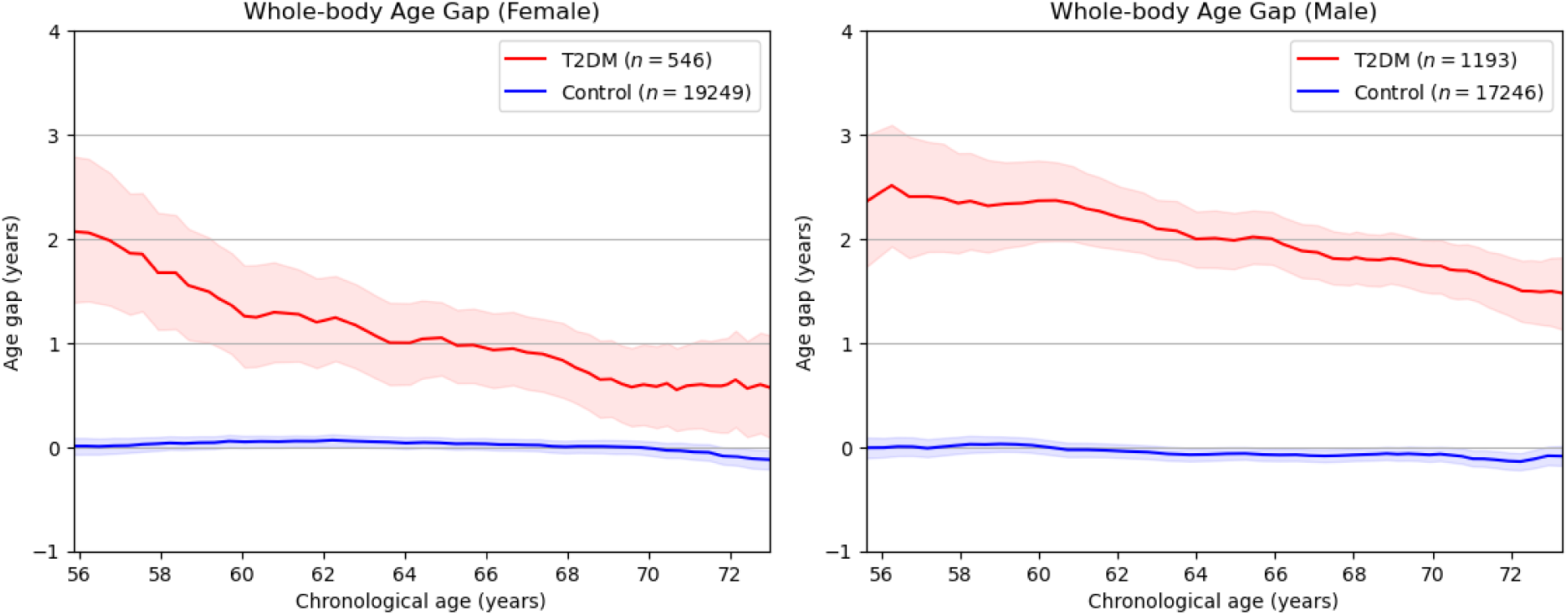
Association of type-2 diabetes (T2DM) and the predicted (and bias-corrected) age gap (age gap = MA - CA), as a function of CA, computed as a sliding window with average bandwidth ± 10 years. The age gap was corrected to a value close to zero for all ages in the control group. The T2DM group exhibits a substantially higher mean age gap for all CAs compared to the controls. Confidence intervals (CIs) were generated by bootstrapping with 1000 samples.

The results of the survival analysis are presented as Kaplan-Meier plots in Fig. 7.

**Figure 7:**
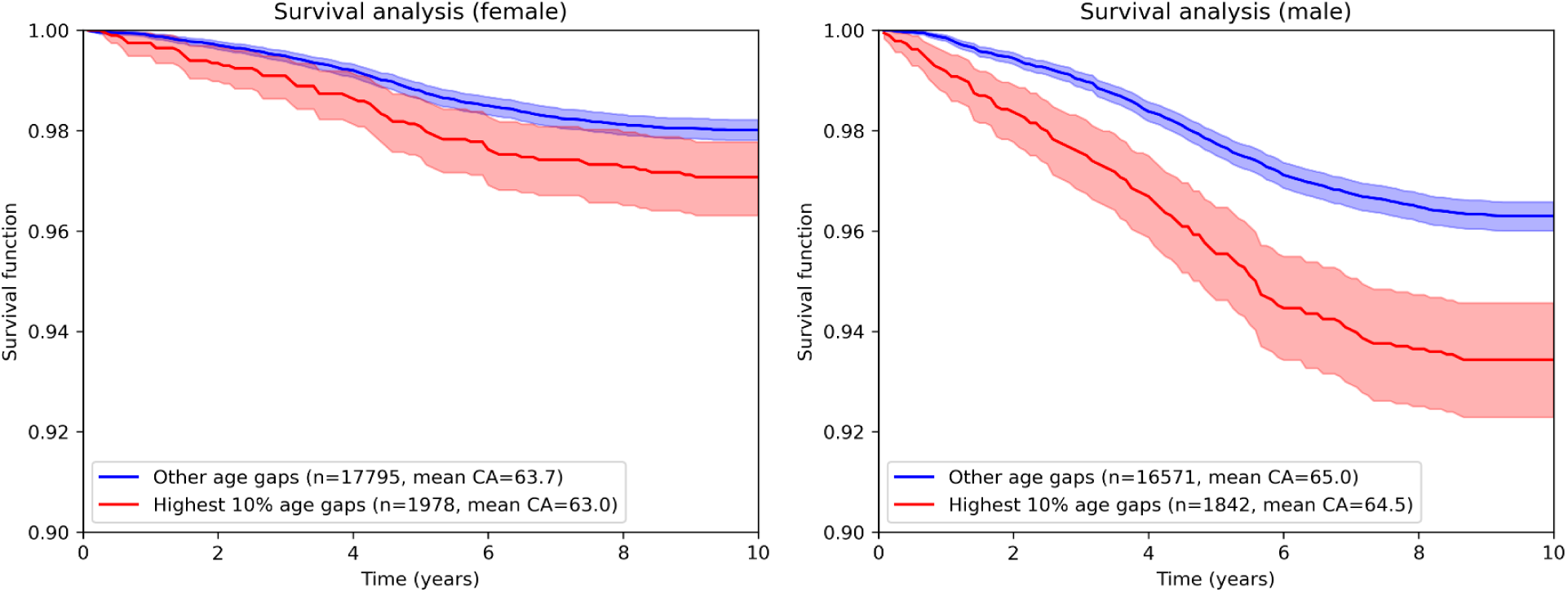
Survival analysis based on the predicted MAs. The highest 10% age gaps denote the group of subjects with an age gap larger than the top 10th percentile in each sub-cohort. The survival function models the number of subjects surviving beyond a given time point. A statistically significant relationship between age gap and survival is observed in both males and females, but is more pronounced in males. A 10-year or longer follow-up time is available for all analyzed subjects. CIs were generated by bootstrapping with 1000 samples.

Subjects with the largest age gap (larger than the 10th percentile) have a significantly higher all-cause mortality compared to the rest of the cohort (adjusted p-value for males: <2.86 * 10^−9^, adjusted p-value for females: < 0.0096).

The prediction of all-cause mortality from CA and MA is presented in Table 3.

**Table 3:** All-cause mortality prediction with logistic regression from CA and MA. The Bonferroni corrected P-values (with correction multiple 2) are presented along with the AUC values for CA and MA.

|  | Subjects | Events | CA AUC | MA AUC | P-value |
| --- | --- | --- | --- | --- | --- |
| Female | 19773 | 252 | 0.698 | 0.705 | 0.479 |
| Male | 18413 | 456 | 0.719 | 0.733 | 0.005 |

We found a statistically significant difference between CA and MA as predictors of all-cause mortality for males (456 events), and no statistically significant difference for females (252 events).

## Discussion

We developed an image-to-biomarker method for tissue-specific spatially standardized predictions (TS-SSP) using supervoxel-based interpretable features and applied it to the prediction of MA from neck-to-knee Dixon MR images in the UKBB, additionally evaluating its clinical relevance in terms of T2DM and all-cause mortality. The method achieved an MAE of 1.951 years on the female cohort (*n=1980*) and an MAE of 2.057 years on the male cohort (*n=1844*), evaluated in comparison to CA, also outperforming features extracted using the state-of-the-art segmentation method VIBESegmentator. To the best of our knowledge, these results represent the highest R^2^ and lowest MAE on age prediction achieved from neck-to-knee MR images. However, we note that, e.g., subset sizes, age ranges, and disease prevalence make direct comparisons to other published results challenging.

The two most similar studies to ours, Langner et al. (4) and Starck et al. (5), also used neck-to-knee MR images from the UKBB, without sex-stratification. Langner et al. utilized 23,000 images in a 10-fold cross-validation setup, and Starck et al. used a training set of 1,536 images. On their designated test sets, Langner et al. and Starck et al. achieved a mean absolute error (MAE) of 2.47 years and 2.57 years, respectively. Both studies conducted saliency analyses using post-hoc explainability methods, aggregating the saliency maps using image registration. The models by Langner et al. predominantly highlighted the aortic arch and the knees, while the model by Starck et al. emphasized the spine, the autochthonous back muscles, and the heart, and its adjacent vessels, all congruent with our findings. In another study, age was predicted from MR brain scans acquired of 15,000 healthy subjects, using a patch-based ensemble approach (6), yielding a MAE of 1.96 years and an R^2^ of 0.87. This performance is comparable to what is achieved in this study, even though we hypothesize that predicting the CA from healthy subjects is a less challenging task than when including subjects with, e.g., cardiometabolic diseases. In a study with chest CT scans as input, with ages between 55 and 75 years, an MAE of 1.84 years was achieved (lower MAE but narrower age range), observing an association between the predicted ages and outcomes in cancer patients (28).

In this study, we have fitted linear models to supervoxel-based standardized imaging biomarkers to predict clinically relevant MAs as well as generate 3D interpretability maps to analyze the model associations. The number of supervoxels can be adjusted with a single parameter, for a trade-off between the number of features and the spatial resolution, and tissue-specific supervoxel features can reduce the effect of mixing of different tissues. In the absence of reference MAs, we relied on a hypothesized capacity of the models to learn the patterns of normal aging from the CAs. We observed that for 3000 PCA components, the CA prediction exhibited very high performance. However, the resulting MAs did not exhibit a strong association with T2DM and mortality. Using a more parsimonious model with the 10 strongest components, the performance on CA prediction was inferior, but the association with T2DM and mortality was stronger and statistically significant, and the resulting interpretability maps were found to be more interpretable. We conclude that the models achieving high CA predictive performance may lose information about pathology and morphological aging, and thereby be less clinically relevant. On the other hand, the models with inferior CA predictive performance can be useful for estimating MA and its association with diseases and all-cause mortality. Future studies need to consider this balance between prediction accuracy and interpretability. In addition, age prediction studies need to consider this well-known challenge of tracking biological age models using CA.

This work has several limitations. Inter-subject registration is challenging due to differences in anatomy and appearance, and the resulting errors and ambiguities may present as feature noise. Bias can be introduced in the selection of the reference scan, although some bias is mitigated by deriving the supervoxels from the mean image. Partial volume effects may still exist within the defined tissue groups. Analysis of the association between, in particular, mean FF and age is complicated by the use of FF to define the tissue groups. A subset of the images contains stitching artifacts and undetected water-fat swaps (i.e., misidentification of water-based tissue as fat-based tissue and vice versa), which may influence both the image registration and feature extraction. For females, both the number of deceased subjects and the number of subjects with T2DM were lower than for males, and the associations between MA and T2DM, as well as all-cause mortality, were substantially weaker than for males.

We have identified numerous avenues for future studies: Replication of the MA prediction and evaluation of its clinical relevance in other cohorts with different population characteristics, prospective testing of the method on new subjects, and estimation of its sensitivity to realistic domain shift. The image-to-biomarker extraction method can easily be adapted to enable studies using subsets of body regions, tissues, organs, and image feature types, enabling a wide range of future medical studies. We aim to evaluate the proposed method on other interpretable precision medicine applications, such as the prediction of future diabetes and cardiovascular disease, and oncological applications, such as the prediction of treatment response, treatment side-effects, and overall survival from CT or PET/CT image data. The supervoxels could be constructed in a more data-driven manner, optimized to maximize performance on the prediction task, and the image registration methodology might be further improved. Future studies could explore additional tissue groups, and tissue grouping being performed in the subject space rather than in reference space, reducing partial volume effects on the tissue group assignment.

## Conclusion

We developed a novel image-to-biomarker conversion methodology and applied it to MA prediction from whole-body MR images in the UKBB. Linear and interpretable predictions were performed using localized and tissue-specific features, resulting in state-of-the-art age prediction performance from whole-body MR images, outperforming a multi-organ and tissue segmentation-based approach and previously reported CNN-based prediction results. The MAs were significantly associated with T2DM and all-cause mortality.

## Data Availability

The data used in this study is from the UK Biobank and can be accessed by researchers after a successful application.

https://www.ukbiobank.ac.uk/

## Acknowledgments and declarations of interest

This research was enabled by grants from the Swedish Heart-Lung Foundation (20240402) and the Swedish Research Council (2016-01040, 2019-04756, 2023-03607), EXODIAB, and Uppsala Diabetes Center (UDC).

This research has been conducted using the UK Biobank Resource under Application Number 14237 and the Swedish National Infrastructure for Computing (SNIC, sens2019016). The computations and data handling were enabled by resources provided by the Swedish National Infrastructure for Computing (SNIC) at Uppsala University, partially funded by the Swedish Research Council through grant agreement no. 2018-05973.

The funding bodies had no influence on the research, preparation of the manuscript, or the decision to publish.

Images from the UK Biobank reproduced by kind permission of UK Biobank ©. This work uses data provided by patients and collected by the NHS as part of their care and support.

Authors J.Ö and J.K declare a patent-pending invention relevant for the proposed method. Authors A.B, Y.U., R. K. G., E.L., and H.A. declare no conflicts of interest. H.A. and J.K. are co-founders and part-time employees of Antaros Medical AB, Mölndal, Sweden.

## Author contributions

J.O.: Writing of the first draft of the manuscript, study design, data analysis, code, method development, statistics, production of figures, and manuscript revision.

A.B.: Writing of the first draft of the manuscript, data analysis, code, production of figures, and manuscript revision.

Y.U.: Data analysis (image registration implementation and processing), and manuscript revision.

E.L.: Study design, statistics, and manuscript revision.

H.A.: Study design, analysis of the interpretability maps, funding acquisition, and manuscript revision.

J.K.: Study design, analysis, method development, production of figures, funding acquisition, and manuscript revision.

## Supplementary material

### A: Image registration

Image registration was performed using a deformable image registration method based on solving sequences of local-block (block-size 12 along all three dimensions) graph-cut optimization problems in a multi-resolution 6-level Gaussian pyramid framework (10, 11, 15). The input consisted of normalized water fraction and fat fraction images, as well as AI-derived masks, obtained with VIBESegmentator, automatically delineating subcutaneous adipose tissue (SAT) and muscle tissue. The channel-wise sum of square differences was used as the objective function for the image registration (both for the intensity images and tissue masks). The water and fat fraction images were given a weight of 1, and the tissue masks were given a weight of 0.6. A spatial regularization term of 0.25 was selected based on preliminary experiments in order to enforce plausibility of the deformations, while still allowing enough flexibility to attain high anatomical overlap. 100 iterations were performed per level in the multi-resolution pyramid (or stopping early in case of no improvement).

The *deform* (*15*) package produced a displacement vector field representing the local voxel-wise correspondences between the reference subject and each of the other subjects. The contents of the individual subject FF volumes were then deformed into the reference space by applying inverse mapping with the displacement vector field to the corresponding FF volume with trilinear interpolation, bringing all FF volumes into the same sex-stratified template space. Additionally, the *deform* package provided the local Jacobian determinant (JD) for each voxel, which is a relative measure of voxel size, where a JD>1 indicates an expansion and a JD<1 indicates a contraction, in comparison to the template subject in that specific voxel, and where JD<0 indicates a fold of space.

### B: Supervoxels

For the SLIC algorithm, the implementation in the scikit-image package (14) was used. The SLIC algorithm implementation is parametrized by the parameter *n_seg_* (fewer than the original number of voxels), controlling the number of supervoxels that the algorithm aims to partition the image into, as illustrated in Fig. 2. A higher *n_seg_* input results in a greater preservation of data with a higher spatial detail and an increased number of features, which in turn increases the noise of the features, the computational time and the size of the feature sets. Moreover, supervoxels are allowed to split or merge during the course of the algorithm’s operation, so the exact number of supervoxels may differ from the provided *n_seg_*.

### C: Tissue-specific feature extraction

Formally, the features are defined as follows. Let *x* denote a voxel in the reference image space, *S* denote a supervoxel defined in the reference image space, and t denote the tissue group (e.g., lean tissue). *I(x*) denotes the FF of voxel *x*, *JD(x)* denotes the Jacobian Determinant of voxel *x*, *T_t_(x)* is a real-valued tissue weight between 0 and 1 for tissue group *t* and voxel *x*, *N_t_(S)* is the total weight of the voxels (also referred to as Voxel Count when used as a feature) in supervoxel *S* for tissue group *t*. The tissue-specific features *f_t_* for tissue group *t* are then given by weighted averages, sums, and standard deviations, given by

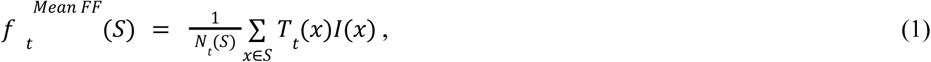

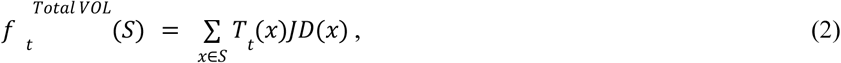

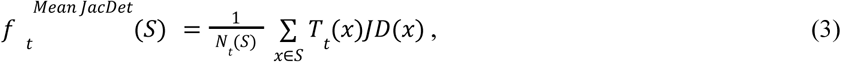

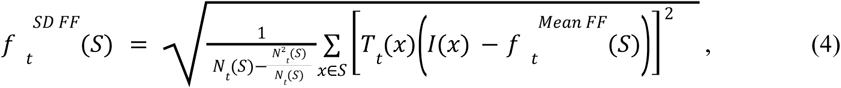

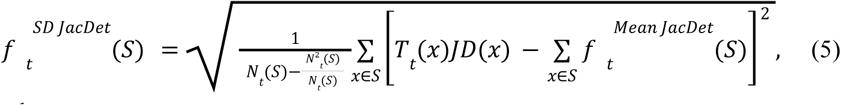

where

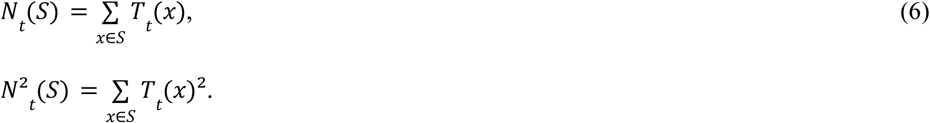

In this study, all tissue weights are binary, i.e., every voxel is assigned to a single tissue group. In general, the features are defined for non-binary tissue weights, thereby supporting the probabilistic assignments of voxels into tissue groups (e.g., a probabilistic model predicts that voxel x contains lean tissue with probability 0.6 and adipose tissue with probability 0.4).

The deformed FF maps were obtained by application of inverse mapping with tri-linear interpolation. JD fields were computed from the image registration deformation fields.

### D: Feature pre-processing: z-score normalization, imputation of missing features, and dimensionality reduction

Given a feature *f_i_*, its mean *f̄*, standard deviation SD(f), and a limit c denoting how many standard deviations to clip the features at, the clipped z-scores were computed by

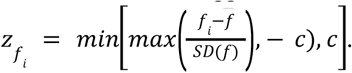

In all experiments, we used c=3 based on preliminary experiments on the CV sets.

When using multiple tissue groups, some supervoxels may not contain enough voxels belonging to a group for the features to be well-defined (e.g., several supervoxels in the liver lacked adipose tissue voxels in all subjects). To account for this, features where 90% or more across the entire training set were undefined were dropped. Undefined features that were not dropped were imputed with a z-score of 0.

### E: Details on the PCA analysis

The number of input feature dimensions was reduced to

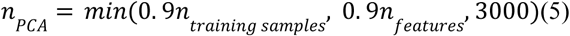

principal components, limiting the number of components to fewer than the theoretical limit, after observing instabilities in the performance without this restriction. When a low number of PCA components is desired in order to limit the flexibility of the regression, we computed the univariate absolute Pearson correlation coefficient between each PCA component and the target variable (training set) and selected the *n_Top–k PCA_* strongest association with the target variable (here, CA). components that exhibit the

The statistics used for z-score normalization and the PCA components were computed during model training from the training set features. The constant *c* was set to 3 (clipping the features at −3 and +3 standard deviations), a decision made from initial tests during cross-validation to obtain a good trade-off between preserving informative feature variation and removing noise and outliers.

### F: Linear modelling

Linear regression models were selected during initial rounds of cross-validation experiments for having several beneficial properties compared to non-linear models, e.g., multi-layer perceptron models: 1) efficiency of model fitting and prediction on unseen data, 2) the same observed prediction performance or better than the deeper models tested, and 3) superior interpretability of the linear models over more complex models.

### G: Debiasing of the MAs for the computation of age gaps

For 10 equidistant grid-points in the range between the minimum and the maximum age, we computed the moving average MA, with weights drawn from an unnormalized Laplace kernel with a bandwidth of 0.25 years, given by

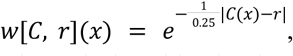

where r is the grid-point (in years), C(x) denotes the CA of subject x. The updated MAs were then given by linear interpolation between the grid points. The correlation before and after bias correction is presented in Table G1.

**Table G1:** Effect of debiasing the 10 and 3000 component MAs. Before debiasing (pre), the correlation was strongly negative, and after debiasing (post), the correlation was near zero.

| Components | Sex | r (Pre) | P-value (Pre) | r (Post) | P-value (Post) |
| --- | --- | --- | --- | --- | --- |
| 10 | Female | -0.607 | 0.0 | -0.0067 | 0.352 |
|  | Male | -0.637 | 0.0 | 0.0096 | 0.195 |
| 3000 | Female | -0.356 | 0.0 | 0.0343 | $1.31 \cdot 10^{-5}$ |
| | Male | -0.358 | 0.0 | 0.0167 | $3.33 \cdot 10^{-6}$ |

### H: Statistics

Bonferroni-adjusted P-values for the comparative tests between the 3-tissue configuration and the 1-tissue configuration, as well as between the 3-tissue configuration with supervoxels and the 3-tissue configuration with VIBESegmentator-derived masks, are presented in Table H1. Bonferroni-adjusted P-values for the comparative tests between the 3-tissue configuration and the 1-tissue configuration of VIBESegmentator-derived masks are presented in Table H2.

**Table H1.** Bonferroni-adjusted P-values for Wilcoxon signed-rank tests on absolute differences of predictions and reference CA between the 9 different models trained on the 9 different folds using cross-validation. The supervoxel experiments were performed with the 5000 supervoxel setup and 3000 PCA components. All adjusted p-values indicate statistically significant differences (threshold 0.05) except for fold 6 for males in the comparison between the 3-tissue configuration and the 1-tissue configuration.

| Method | Sex | 1 | 2 | 3 | 4 | 5 | 6 | 7 | 8 | 9 |
| --- | --- | --- | --- | --- | --- | --- | --- | --- | --- | --- |
| 3-T SV and 1-T SV | Female | $8.92 \cdot 10^{-5}$ | $9.87 \cdot 10^{-6}$ | $1.15 \cdot 10^{-5}$ | $1.74 \cdot 10^{-4}$ | $2.25 \cdot 10^{-5}$ | $4.63 \cdot 10^{-6}$ | $4.83 \cdot 10^{-5}$ | $3.82 \cdot 10^{-3}$ | $2.84 \cdot 10^{-4}$ |
| | Male | $7.05 \cdot 10^{-5}$ | $9.60 \cdot 10^{-5}$ | $1.66 \cdot 10^{-3}$ | $1.89 \cdot 10^{-4}$ | $2.72 \cdot 10^{-4}$ | $9.02 \cdot 10^{-1}$ | $8.70 \cdot 10^{-3}$ | $2.75 \cdot 10^{-2}$ | $1.15 \cdot 10^{-3}$ |
| 3-T SV and 3-T VIBESeg. | Female | $4.10 \cdot 10^{-88}$ | $1.85 \cdot 10^{-87}$ | $1.20 \cdot 10^{-89}$ | $6.98 \cdot 10^{-86}$ | $2.15 \cdot 10^{-88}$ | $1.40 \cdot 10^{-87}$ | $6.38 \cdot 10^{-85}$ | $5.88 \cdot 10^{-84}$ | $2.88 \cdot 10^{-85}$ |
| | Male | $2.49 \cdot 10^{-63}$ | $3.18 \cdot 10^{-64}$ | $9.66 \cdot 10^{-60}$ | $2.71 \cdot 10^{-66}$ | $3.78 \cdot 10^{-62}$ | $1.09 \cdot 10^{-59}$ | $7.12 \cdot 10^{-62}$ | $6.49 \cdot 10^{-64}$ | $4.64 \cdot 10^{-62}$ |

**Table H2.** Bonferroni-adjusted P-values for Wilcoxon signed-rank tests on absolute differences of predictions and reference CA between the 9 different models trained on the 9 different folds using cross-validation. All adjusted p-values indicate statistically significant differences (threshold 0.05).

| Method | Sex | 1 | 2 | 3 | 4 | 5 | 6 | 7 | 8 | 9 |
| --- | --- | --- | --- | --- | --- | --- | --- | --- | --- | --- |
| 3-T VIBESeg.<br>and<br>1-T VIBESeg. | Female | $2.83 \times 10^{-5}$ | $4.26 \times 10^{-6}$ | $4.00 \times 10^{-5}$ | $2.84 \times 10^{-6}$ | $4.81 \times 10^{-6}$ | $4.49 \times 10^{-5}$ | $6.68 \times 10^{-6}$ | $7.64 \times 10^{-5}$ | $8.71 \times 10^{-6}$ |
| | Male | $7.06 \times 10^{-6}$ | $4.83 \times 10^{-7}$ | $2.13 \times 10^{-6}$ | $1.68 \times 10^{-5}$ | $7.50 \times 10^{-7}$ | $1.01 \times 10^{-6}$ | $3.13 \times 10^{-6}$ | $1.60 \times 10^{-5}$ | $2.26 \times 10^{-6}$ |

### I: Ablation studies

In Fig. I1, the cross-validation performance on age prediction as a function of the number of training subjects is presented.

**Figure I1:**
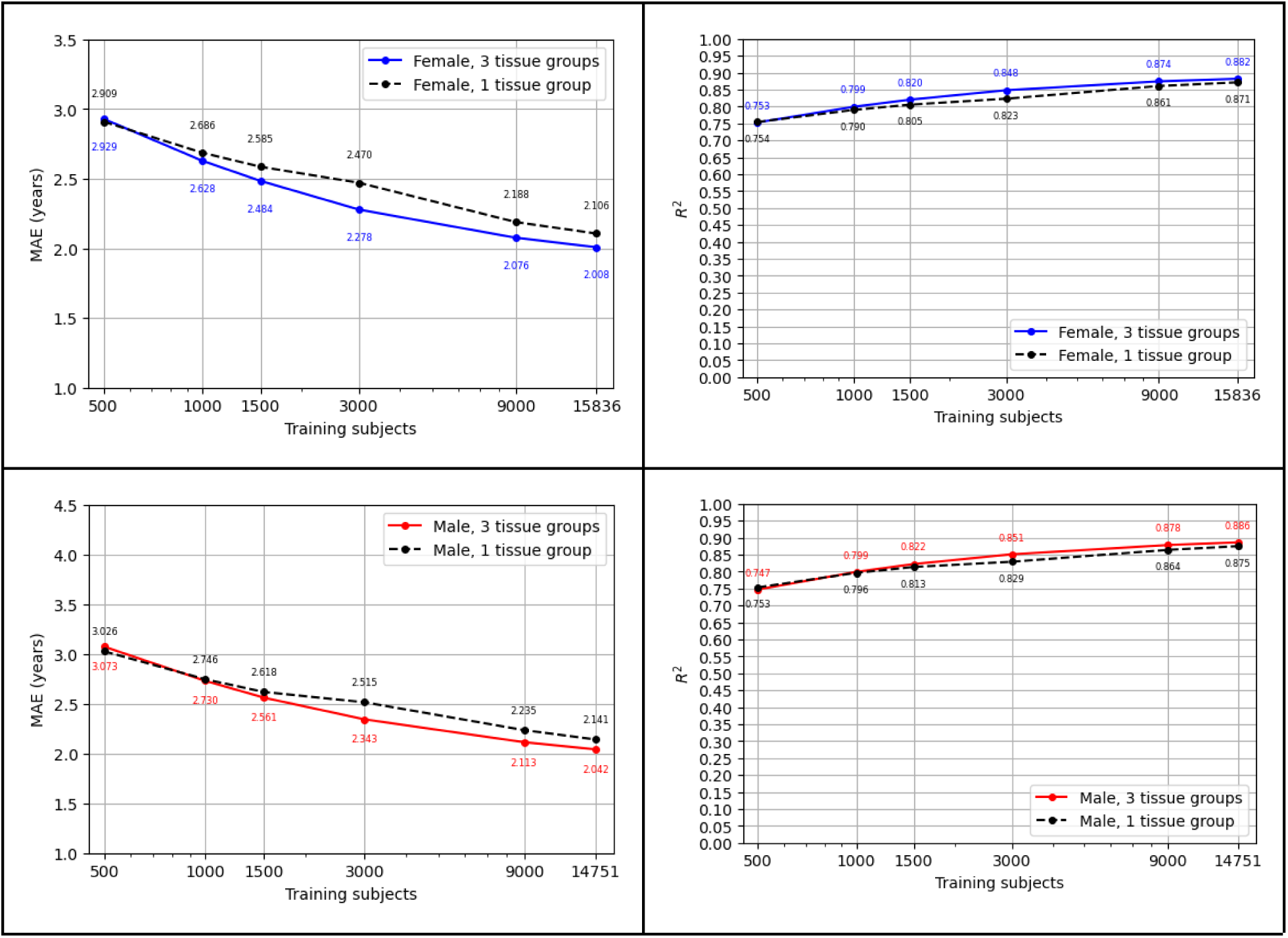
The effect of different numbers of training subjects on R^2^ and MAE performance on age prediction across the cross-validation, comparing the 1-tissue group configuration with the 3-tissue group configuration. A higher R^2^ and a lower MAE indicate better performance. The y-axis is scaled linearly, while the x-axis is scaled logarithmically. 5000 supervoxels were used for these experiments.

### J: Chronological age prediction performance with 10 PCA components vs 3000 PCA components

Table J1 presents a comparison of the performance of CA prediction with 10 PCA components and 3000 PCA components.

**Table J1:**
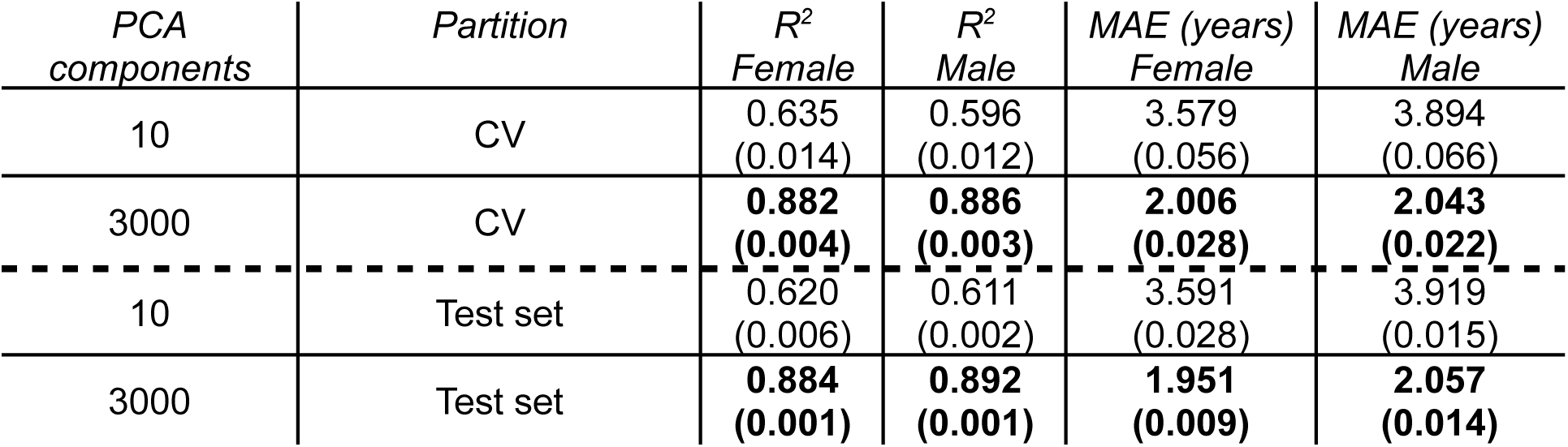
Performance of prediction of CA, evaluated with 10 and 3000 PCA components. The performance was measured using R^2^ and MAE. Each cell presents the mean results of nine cross-validation models on the test set, with the standard deviation across models in parentheses.

| *PCA components* | *Partition* | *$R^2$*<br>*Female* | *$R^2$*<br>*Male* | *MAE (years)*<br>*Female* | *MAE (years)*<br>*Male* |
| --- | --- | --- | --- | --- | --- |
| 10 | CV | 0.635<br>(0.014) | 0.596<br>(0.012) | 3.579<br>(0.056) | 3.894<br>(0.066) |
| 3000 | CV | **0.882**<br>**(0.004)** | **0.886**<br>**(0.003)** | **2.006**<br>**(0.028)** | **2.043**<br>**(0.022)** |
| 10 | Test set | 0.620<br>(0.006) | 0.611<br>(0.002) | 3.591<br>(0.028) | 3.919<br>(0.015) |
| 3000 | Test set | **0.884**<br>**(0.001)** | **0.892**<br>**(0.001)** | **1.951**<br>**(0.009)** | **2.057**<br>**(0.014)** |

### K: Association of the predicted MAs with diabetes and all-cause mortality

The results of the association of the age gap and prevalence of T2DM are presented in Fig. K1.

**Fig K1:**
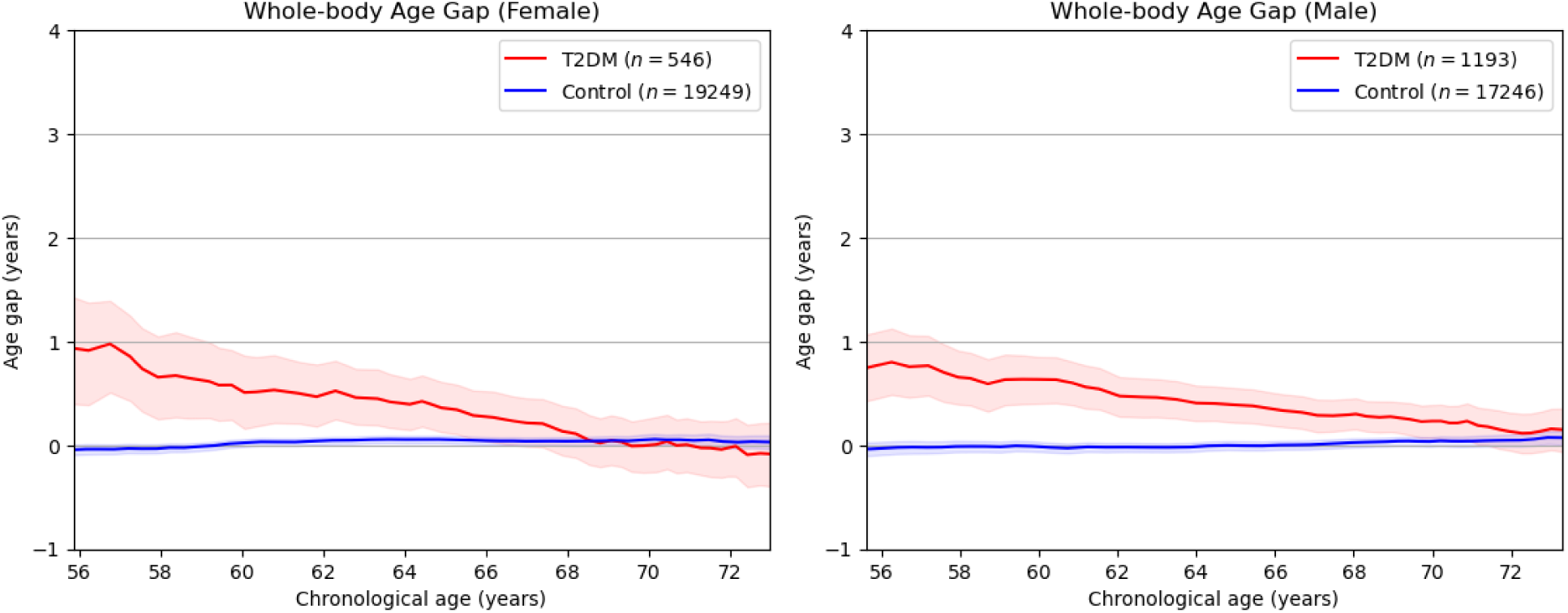
Association of type-2 diabetes (T2DM) and the predicted (and bias-corrected) MAs from 3000 PCA components (compared to 10 for the results in the main text), as a function of CA, computed as a sliding window with average bandwidth ± 10 years. The age gap was corrected to a value close to zero for all ages in the control group. The T2DM group exhibits a substantially higher mean age gap for all CAs. CIs were generated by bootstrapping with 1000 samples.

The results of the survival analysis for 3000 PCA components are presented as Kaplan-Meier plots in Fig. K2.

**Fig K2:**
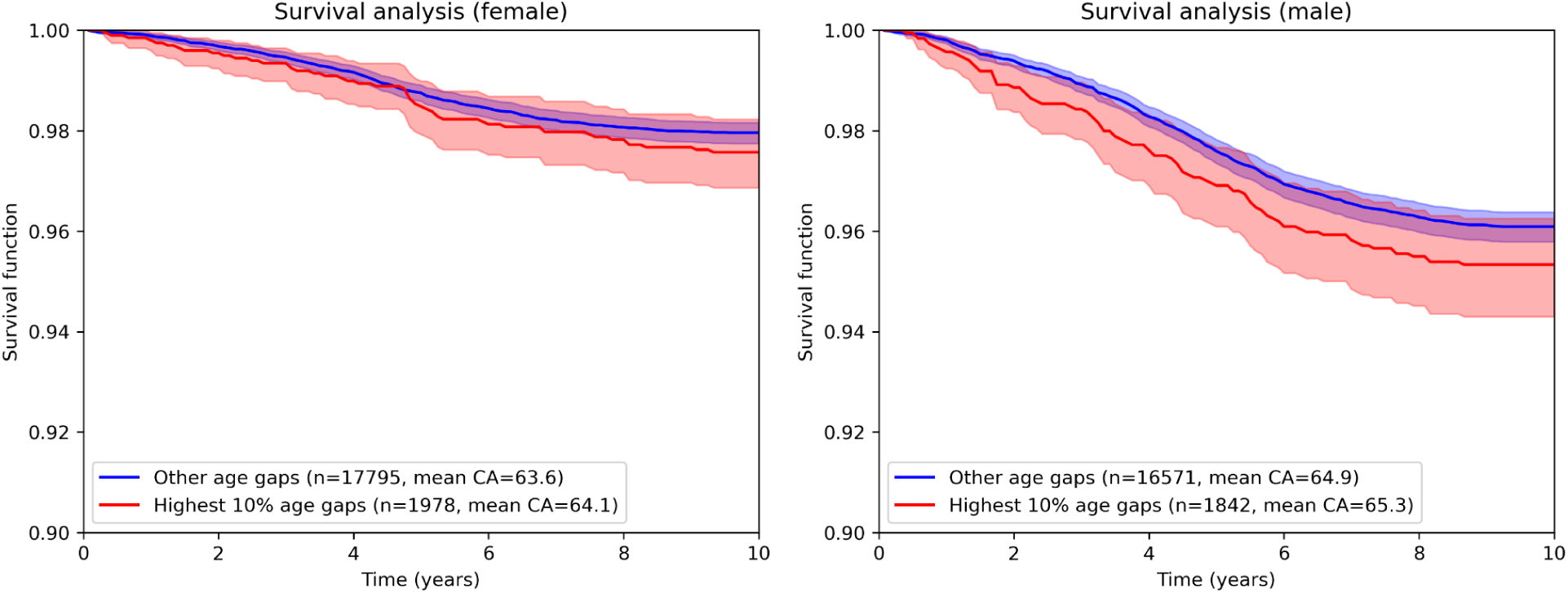
Survival analysis based on the predicted MAs for the model with 3000 PCA components. The highest 10% age gaps denote the group of subjects with an age gap larger than the top 10th percentile in each sub-cohort. The survival function models the number of subjects surviving beyond a given time point. A 10-year or longer follow-up time is available for all analyzed subjects. CIs were generated by bootstrapping with 1000 samples.

### L: Software

*SimpleITK*, v2.3.1 (18), *NumPy*, v1.26.4 (S1), *scikit-image*, v0.22.0 (S2), *scikit-learn*, v1.2.2 (S3), and *deform,* v0.5.2 (10, 11, 15) (S4) with Python 3.8.8 were the main software packages and versions used for the image data analysis. VIBESegmentator model 80 was used for segmentation.

### M: Tissue-wise and feature-wise age prediction performance

| Female | Quality measure | Mean FF | Total Volume | Mean JacDet | Voxel Count | SD FF | SD JacDet |
| --- | --- | --- | --- | --- | --- | --- | --- |
| Lean tissue | MAE (y) | 2.930<br>(0.012) | 2.674<br>(0.0014) | 2.758<br>(0.016) | 2.870<br>(0.021) | 3.123<br>(0.010) | 3.143<br>(0.024) |
|  | R2 | 0.749<br>(0.002) | 0.788<br>(0.002) | 0.773<br>(0.002) | 0.754<br>(0.003) | 0.711<br>(0.002) | 0.705<br>(0.004) |
| Medium FF tissue | MAE (y) | 2.557<br>(0.014) | 2.495<br>(0.016) | 2.614<br>(0.017) | 2.474<br>(0.013) | 2.651<br>(0.008) | 2.878<br>(0.023) |
|  | R2 | 0.805<br>(0.001) | 0.815<br>(0.002) | 0.793<br>(0.002) | 0.816<br>(0.002) | 0.791<br>(0.001) | 0.752<br>(0.004) |
| Adipose tissue | MAE (y) | 3.038<br>(0.022) | 2.926<br>(0.020) | 2.945<br>(0.015) | 2.805<br>(0.015) | 3.180<br>(0.012) | 3.192<br>(0.014) |
|  | R2 | 0.726<br>(0.004) | 0.741<br>(0.002) | 0.741<br>(0.003) | 0.765<br>(0.003) | 0.698<br>(0.002) | 0.702<br>(0.003) |

| Male | Quality measure | Mean FF | Total Volume | Mean JacDet | Voxel Count | SD FF | SD JacDet |
| --- | --- | --- | --- | --- | --- | --- | --- |
| Lean tissue | MAE (y) | 2.970<br>(0.016) | 2.673<br>(0.012) | 2.794<br>(0.012) | 2.942<br>(0.013) | 3.060<br>(0.011) | 3.188<br>(0.023) |
|  | R2 | 0.773<br>(0.001) | 0.816<br>(0.001) | 0.794<br>(0.002) | 0.778<br>(0.002) | 0.760<br>(0.002) | 0.735<br>(0.003) |
| Medium FF tissue | MAE (y) | 2.690<br>(0.018) | 2.655<br>(0.016) | 2.724<br>(0.010) | 2.553<br>(0.019) | 2.759<br>(0.023) | 3.011<br>(0.021) |
|  | R2 | 0.813<br>(0.002) | 0.816<br>(0.002) | 0.810<br>(0.001) | 0.831<br>(0.002) | 0.799<br>(0.003) | 0.765<br>(0.003) |
| Adipose tissue | MAE (y) | 3.199<br>(0.019) | 3.224<br>(0.013) | 3.162<br>(0.019) | 3.022<br>(0.018) | 3.453<br>(0.024) | 3.488<br>(0.015) |
|  | R2 | 0.734<br>(0.003) | 0.732<br>(0.002) | 0.745<br>(0.003) | 0.758<br>(0.002) | 0.690<br>(0.004) | 0.687<br>(0.003) |

### N: Computational pipeline and software runtime

The main steps performed by the analysis pipeline from raw images are sequentially in order:

1. Stitching of the images from the different MR imaging stations.
2. Computation of the FF, WF, and sum images.
3. Segmentation using VIBESegmentator using the sum images as input.
4. Registration of the image to the reference image.
5. Computation of the deformed FF image and the JacDet image.
6. Tissue group assignment based on deformed FF images.
7. Supervoxel-based feature extraction by supervoxel, tissue group, and feature combination.
8. Z-score standardization and filtering of features.
9. PCA dimensionality reduction.
10. Application of the linear model to the PCA-components.

The approximate runtime of the full pipeline on an Intel i9-13900K, 64 GB RAM machine for a new subject is 5 minutes. The majority of the runtime is due to the inter-subject image registration, and to a lesser part, the stitching, segmentation, and feature extraction. The PCA-based dimensionality reduction and application of the linear model are almost a negligible part of the runtime.

### O: Interpretability studies and interpretability maps

The interpretability studies involve the creation of visual interpretability maps, showing the relationships between the features and the target variable used by the models, overlayed on the standardized supervoxels in the reference image space where they were computed. Starting with the coefficients of the linear models taking the PCA-components as inputs, we map back their relationship to the original input features, by multiplication of the PCA loadings with the model coefficients, as given by

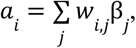

where *w_i,j_*denotes the PCA loadings for feature i and component *j*, β*_j_* denotes the linear coefficient from PCA component *j* and the target variable.

For visualization, these feature coefficients are normalized to the −1 to +1 range by division by the maximum absolute value over all *a_i_*.

To simplify the viewing of the interpretability maps, each association *g_i_* is plotted in the supervoxel it was computed from, and only in the voxels belonging to the tissue group corresponding to feature *i* in the reference subject. This enables the simultaneous viewing of the associations from all tissue groups in a single map.

